# Factors Influencing Job Satisfaction among Healthcare Providers in a Specialty Department: a cross-sectional study

**DOI:** 10.1101/2023.06.24.23291811

**Authors:** Tanvir Haider, Syeda Sumaiya Efa, Md. Emam Hossain, Shamima Gulshan Ara Shampa, Syed Nafi Mahdee, Rafsan Reza, Md. Fuad Al Fidah

## Abstract

**Background:** Job satisfaction of professionals affects health, advancement, performance, and development, as well as the institution, employer, or organization. Healthcare professionals who are satisfied with their job have a higher probability of delivering excellent healthcare. In Bangladesh, the challenges of public health highlight the importance of having a competent healthcare workforce to provide an improved quality healthcare service.

**Objectives:** The objective of this study was to evaluate the degree of job satisfaction and identify the factors that contribute to it among healthcare providers employed in the BMT unit of DMCH.

**Methods:** This cross-sectional study was conducted for the period of six months, from July to December 2014. The study population was all the healthcare professionals at the BMT unit of DMCH, Bangladesh. and consisted of doctors (n=20), nurses (n=15), and laboratory technicians (n=5). A semi structured self-administered questionnaire was used to collect the preliminary data. All ethical issues were maintained strictly.

**Results:** Most (95.0%) of the study respondents were at or below the age of 40 years with a mean (±SD) of 30.1 (±7.92) years. Most of them were female (52.5%), and lab technician, nurse and doctors had a frequency of 12.5%, 37.5%, and 50.0% respectively. Among the study participants, 65.0% were satisfied with their jobs, 35% were dissatisfied. Statistically significant association was found between sex (p=0.011) and profession (p < 0.001) with level of job satisfaction among respondents.

**Conclusion:** Job satisfaction is important for healthcare professionals, patients, and institutions. Satisfied professionals provide better care, while low job satisfaction leads to turnover and decreased access to care. Individual, job-related, and workplace factors influence job satisfaction. This study evaluated job satisfaction among healthcare providers in Bangladesh and identified contributing factors to inform interventions.

## Introduction

Job satisfaction of professionals affects health, advancement, performance, and development, as well as the institution, employer, or organization.^1^ Behavioral and social science studies suggest that job contentment and performance are inseparably connected.^2^ Employee’s job satisfaction is affected by the nature of job. Independence, a clear purpose and outcome, relevance to their role, and regular feedback make employee’s happier. It can be said that organizational stress lowers job satisfaction considerably.^3^ Research conducted by in 1911 was the inspiration for the development of both the idea of job satisfaction and the method of measuring it.^4^ Followed by, a research on the degree of job satisfaction among healthcare professionals in the United States concentrated on laboratory staff in 1971.^5^ Since then, a number of studies on Job satisfaction of healthcare providers (HCP) have been carried out in different parts of the world. Overall health and well-being of the organization as well its employee heavily rely on job satisfaction.^6^

Studies have revealed that healthcare professionals who are satisfied with their job, and work in favorable environments have a higher probability of delivering excellent healthcare,^7^ resulting in increased patient satisfaction^8,9,10,11^ and improved adherence to medical instructions.^12^ Consequently, it helps in the retention of healthcare professionals within the field.^13,14^ Low levels of job satisfaction among health workers can lead to higher turnover rates, which in turn can disrupt the delivery of treatment and limit access to healthcare. Various individual factors, including age, gender, marital status, and years of work experience, can influence job satisfaction. Similarly, job-related factors such as specialization, patient interactions, and work engagement can also impact overall job satisfaction.^15^ Additional elements of the job, including job description, job stability, and income level, as well as characteristics of the workplace, such as the facility type, management style, opportunities for professional growth, teamwork, and availability of resources, can also have an influence. These factors may be associated with the overall work environment.**Error! Bookmark not defined**.

In Bangladesh, the challenges of public health highlight the importance of having a competent healthcare workforce to provide an improved quality healthcare service. Ensuring uniform and high-quality training of the healthcare providers is crucial for attaining excellent medical care.^16^ Unfortunately in Bangladesh, there is not enough HCPs to meet the demand, and the working environment is difficult. This causes a strain in the job satisfaction, and also makes it impossible to provide high-quality medical care.

A historical landmark was achieved with the introduction of the first Bone-Marrow Transplant (BMT) unit by Dhaka Medical College Hospital (DMCH) in March 2014.^17^ A total number of 10 patients underwent transplantation since its journey in 2014. However, with proper logistics and machineries, the Unit is capable of conducting almost double the number of BMTs.^18^ DMCH which is the oldest and most well-known tertiary level hospital in the country is under the jurisdiction of the Government of Bangladesh (GOB). HCPs working in this department undergo vigorous training, are stationed there as per government decree. BMT unit provide care to the patients, notably with AML (Acute Myeloid Leukaemaia) and Multiple Myeloma.

In Bangladesh, annually about 200,000 new cancer cases are added each year and 150,000 suffer death.^19^ Considering the advanced nature of the BMT unit at DMCH and the extensive duties carried out by doctors, nurses, and other healthcare personnel, one can easily comprehend the frequency with which these medical professionals encounter various stressful circumstances in fulfilling their professional obligations. The objective of this study was to evaluate the degree of job satisfaction and identify the factors that contribute to it among healthcare providers employed in the BMT unit of DMCH. The findings of this study will aid in devising appropriate interventions to improve job satisfaction and subsequently enhance work productivity among healthcare professionals in the future.

## Materials and Methods

### Study settings and subjects

This cross-sectional study was conducted for the period of six months, from July to December 2014. The study population was all the healthcare professionals at the BMT unit of DMCH, Bangladesh.

### Sample size and sampling technique

The sample size of the study was 40. All the HCPs from the BMT unit were included in the study, and consisted of doctors (n=20), nurses (n=15), and laboratory technicians (n=5).

### Data collection method and instruments

A semi structured self-administered questionnaire was used to collect the preliminary data. Data was collected by the authors.

### Measurement

#### Level of job satisfaction

A single item was used to measure the outcome variable, i.e. study participants’ level of job satisfaction (What is your level of job satisfaction?). It was measured as a binary variable (1=Dissatisfied, 2=Satisfied).

### Data management

Data quality assurance measures were implemented at both the field and central levels to ensure the accuracy and reliability of the data. The principal investigator had control over the safekeeping of the data. Rigorous checks were conducted to verify the relevance and consistency of the collected data. Any incomplete or missing data was identified, resolved, and verified. The data were coded, categorized, cleaned, and subsequently entered into jamovi version 2.3.26 for data analysis.^20^

### Statistical methods

Data for individual variables were summarized using frequency distribution and focused on mean and standard deviation, while findings was presented in percentage. The relationships between respondents’ socio-demographic characteristics and level of satisfaction were analyzed using Chi-squared test. Normality of two continuous variables (monthly income and duration of training) was tested via the Shapiro-Wilk test and Non-parametric test (Mann-Whitney U test) was used where applicable. A p-value <0.05 was considered statistically significant. All statistical tests were two-sided and performed at a significance level of α=0.05.

## Results

In general, most (95.0%) of the study respondents were at or below the age of 40 years and the rest (5.0%) fell above the age of 40 years with a mean (±SD) of 30.1 (±7.92) years. Most of the study participants were female (52.5%) with, and 47.5% were male healthcare providers. Regarding profession, lab technician, nurse and doctors had a frequency of 12.5%, 37.5%, and 50.0% respectively. Based on the level of education, most of the study participants were graduates (55.0%) followed by the postgraduate and diploma health workers of 30.0% and 15.0% respectively. Most of the healthcare workers (57.5%) of BMT unit had monthly income at or below 20,000 BDT while 37.5% had an income ranging 21,000-30,000 BDT and only a 5.0% had income at or above 31,000 BDT with a mean (±SD) of 22175 (±4248) Tk. Most of the healthcare providers (72.5%) had work experience at or above 9 months, while 27.5% had experience at or below 8 months. Most of the HCPs (62.5%) received national training, followed by local training (37.5%). The training lasted for a mean (±SD) of 4.15 (±1.94) months. Among the study participants, 65.0% were satisfied with their jobs, 35% were dissatisfied (Table 1).

**Table 1:**
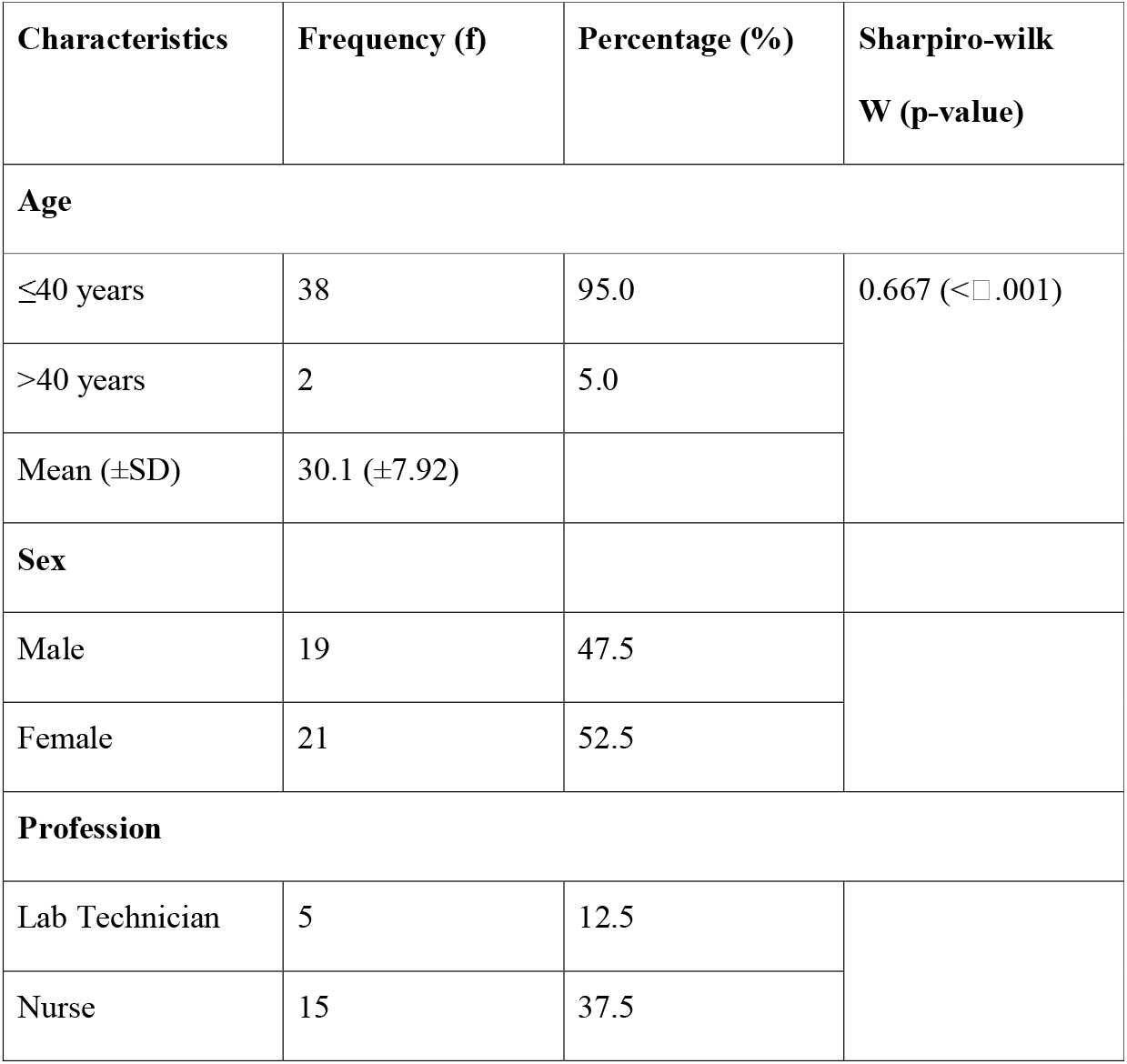
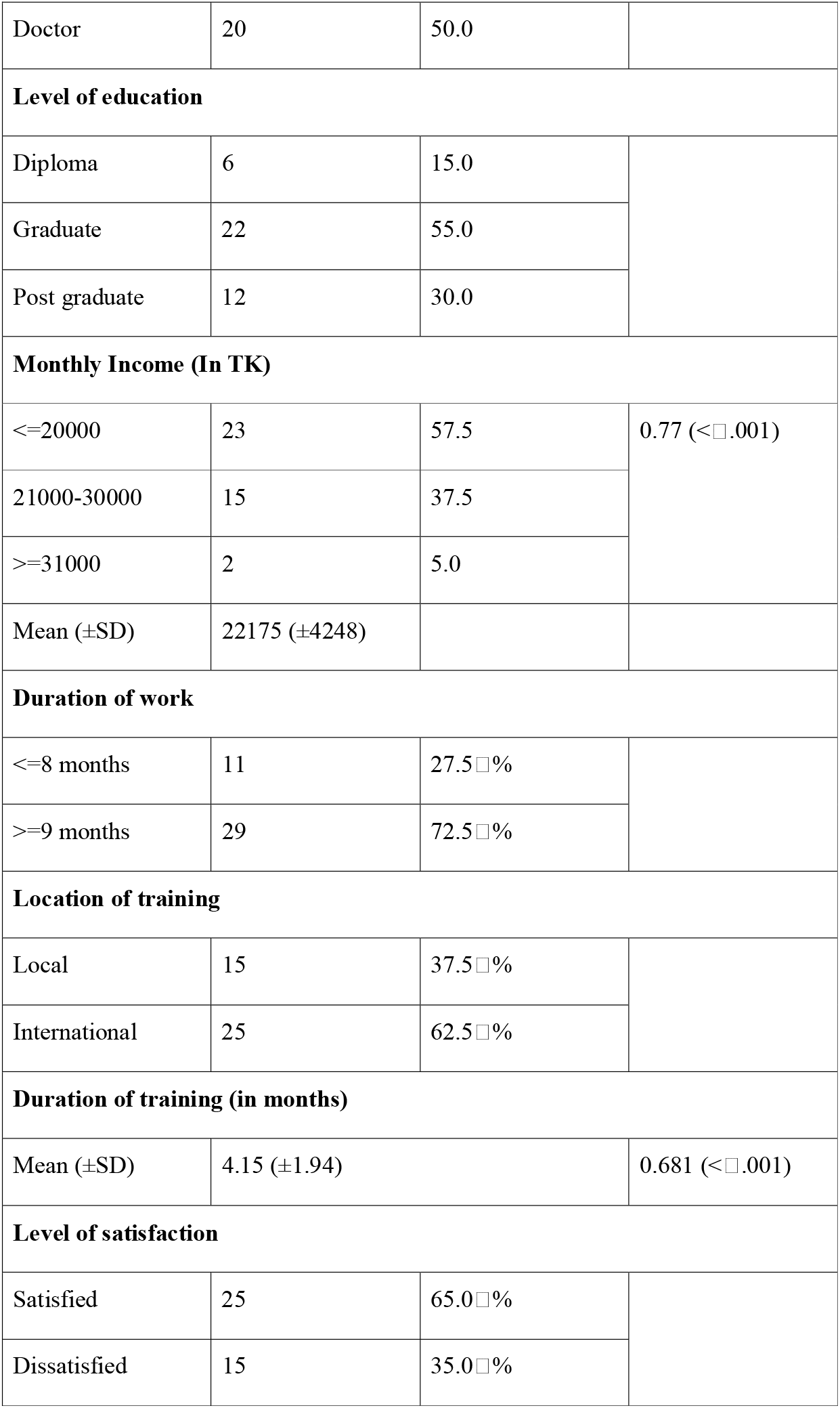
Demographic characteristics of healthcare providers working at BMT unit of DMCH (n=40)

Mann-Whitney U test was conducted to examine the differences on monthly income and duration of training according to the level of job satisfaction. Significant differences were not found among the two categories of participants (Dissatisfied and satisfied) (Table 2).

**Table 2:**
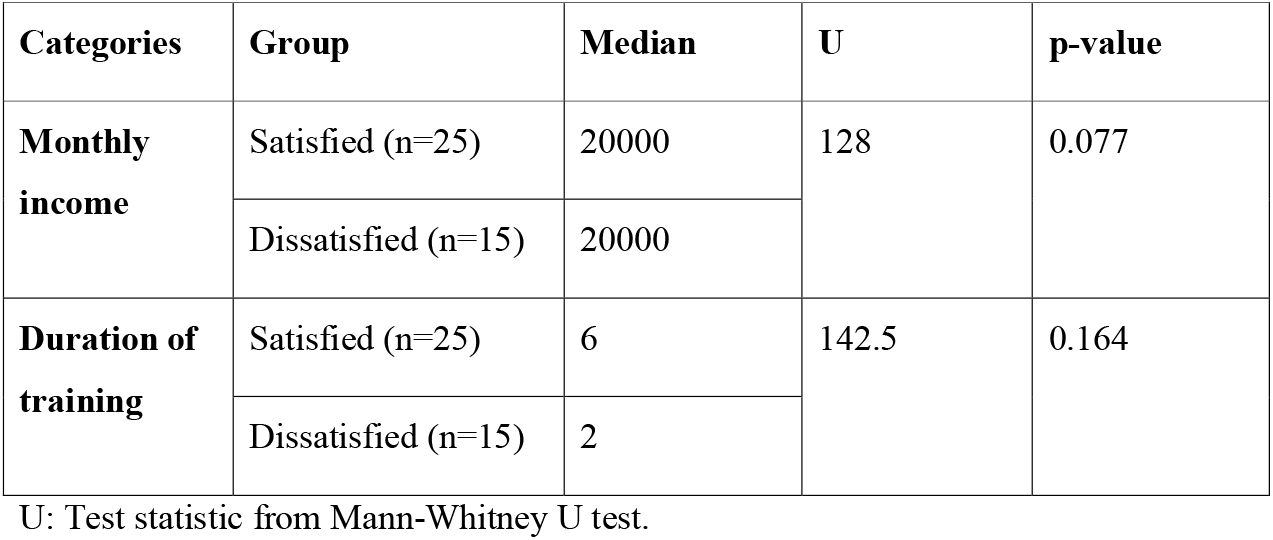
Association between monthly income, duration of training and duration of training with level of job satisfaction.

| Categories | Group | Median | U | p-value |
| --- | --- | --- | --- | --- |
| **Monthly income** | Satisfied (n=25) | 20000 | 128 | 0.077 |
|  | Dissatisfied (n=15) | 20000 |  |  |
| **Duration of training** | Satisfied (n=25) | 6 | 142.5 | 0.164 |
|  | Dissatisfied (n=15) | 2 |  |  |

U: Test statistic from Mann-Whitney U test.

A chi-square test of significance was used to examine the association between selected characteristics and level of job satisfaction. Significant association was found between sex (p=0.011) and profession (p < 0.001) with level of job satisfaction among respondents. However, no statistically significant association was found between age, level of education, duration of work, location of training with level of job satisfaction (Table 3).

**Table 3:** Association between selected independent variables and level of job satisfaction.

| Characteristics | Level of satisfaction |  | p-value |
| --- | --- | --- | --- |
|  | Satisfied | Dissatisfied |  |
| **Age** |  |  | 1.000* |
| ≤40 years | 24 (96□ %) | 14 (93□ %) |  |
| >40 years | 1 (4□ %) | 1 (7□ %) |  |
| **Sex** |  |  | 0.011 |
| Male | 8 (32 □ %) | 11 (73 □ %) |  |
| Female | 17 (68 □ %) | 4 (27 □ %) |  |
| **Profession** |  |  | <0.001* |
| Lab Technician | 3 (12 □ %) | 2 (13 □ %) |  |
| Nurse | 15 (60 □ %) | 0 (0 □ %) |  |
| Doctor | 7 (28 □ %) | 13 (87 □ %) |  |
| **Level of education** |  |  | 0.668* |
| Diploma | 4 (16 □ %) | 2 (13 □ %) |  |
| Graduate | 15 (60 □ %) | 7 (47 □ %) |  |
| Post graduate | 6 (24 □ %) | 6 (40 □ %) |  |
| **Duration of work** |  |  | 0.065* |
| ≤ 8 months | 4 (16 □ %) | 7 (47 □ %) |  |
| ≥ 9 months | 21 (84 □ %) | 8 (53 □ %) |  |
| **Location of training** |  |  | 0.354 |
| Local | 8 (32 □ %) | 7 (47 □ %) |  |
| International | 17 (68 □ %) | 8 (53 □ %) |  |

p\* = p-value from Fisher's exact test.

## Discussion

The study aimed to explore the level of job satisfaction among healthcare providers in a specialized unit of a tertiary level hospital in Bangladesh along with associated factors that may have an impact on the level of job satisfaction. It was found that, 65% of the participants were satisfied in their current position, and 35% was were dissatisfied.

A study conducted in the USA found that, majority of the participants are satisfied with their jobs (64%) and only 7% are dissatisfied. This finding corresponds to the findings of the current study in terms of satisfaction.^21^ Another study conducted in Bangladesh among the Healthcare Professionals of Combined Military Hospitals also reported similar findings.^22^ One reason may be that despite the pressure of the job, working and helping patients with serious illness often provides a sense of achievement.

In our present study, we found that job satisfaction is higher in younger age group (<40 years aged). It may be due to the higher probabilities of attaining opportunities at mid age than that of the old age. However, the study findings contradict the finding of another study conducted in Bangladesh on two leading specialized private hospitals.^23^ This contradiction may arise due to contextual difference as our study was conducted among government employees. HCPs from private hospitals earn more in the start of their carrier than their government counterparts.

Female participants showed higher satisfaction level than their male counterpart, which was found to be statistically significant (p=0.005). It may be due to, traditionally in our country, women are less likely to change work and are satisfied with the minimum requirement.^23^

Regarding educational level, most of the participants were graduates which corresponds to a study conducted in the endoscopy unit in Korea.^24^ The reason behind this may be due to the fact that, in our country, the ratio of doctors, nurses and technicians heavily favors the nurses (41:51:7),^25^ followed by doctors. Government doctors and nurses are, in general, graduates. Hence, the participants of this study are mostly graduates.

The study had several limitations. The sample size was small and thus the result of the study can’t be generalized. Additionally, the cross-sectional nature of the study meant that causal association between independent and dependent variables can not be ascertained. Moreover, no scale was used to measure the level of job satisfaction; rather, a single item question was used to measure job satisfaction.

The study is also with some strengths. As BMT was a relatively new procedure during the time of the study, it provides valuable insights on the job satisfaction among the first batch of HCPs working in such a unit. Also, the study enrolled lab technicians, nurses and doctors to paint a complete picture of the scenario.

## Conclusion

Job satisfaction is important for healthcare professionals, patients, and institutions. Satisfied professionals provide better care, while low job satisfaction leads to turnover and decreased access to care. Individual, job-related, and workplace factors influence job satisfaction. This study evaluated job satisfaction among healthcare providers in Bangladesh and identified contributing factors to inform interventions.

## Data Availability

All data produced in the present study are available upon reasonable request to the authors

## Acknowledgement

The authors are indebted to all study participants for their participation.

